# Potential impact of changes in influenza vaccination coverage and effectiveness on pediatric influenza burden in the United States, 2022/23–2024/25

**DOI:** 10.64898/2026.09.25.26363797

**Authors:** Sinead E. Morris, Sarabeth M. Mathis, Jessie R. Chung, Jennifer DeCuir, Samantha M. Olson, Sascha Ellington, Alissa O’Halloran, Dawud Ujamaa, Catherine H. Bozio, Anurag Jain, Anup Srivastav, Peng-jun Lu, Lisa A. Grohskopf, Carrie Reed, A. Danielle Iuliano, Matthew Biggerstaff

## Abstract

**Background and Objectives:** Influenza vaccination has a strong safety record and is effective in reducing pediatric influenza-associated morbidity and mortality. However, pediatric vaccination coverage (VC) has declined since the 2019/20 influenza season in the United States. We estimated the impact of increased vaccine protection (through increased VC and/or vaccine effectiveness [VE]) on pediatric influenza-associated burden during the 2022/23–2024/25 seasons.

**Methods:** We used a compartmental model and laboratory-confirmed influenza-associated hospitalization, VC, and VE data among children 6 months–17 years to estimate influenza burden from 2022/23–2024/25 (the ‘baseline’ scenario) and the impact of vaccination. We simulated scenarios that (1) increased VC by five percentage points; (2) increased VE by five percentage points; (3) increased VC and VE by five percentage points; and (4) increased VC to levels achieved in 2019/20 (the season with the highest pediatric VC to date). We estimated the difference in hospitalizations and deaths between each scenario and the baseline scenario.

**Results:** Among children 6 months–17 years, an estimated 120,000 influenza-associated hospitalizations and 1,650 deaths occurred across all seasons at baseline. Increasing VC to 2019/20 levels had the greatest estimated impact, reducing hospitalizations by 8,670 (7.2%) and deaths by 130 (7.9%). Increasing both VC and VE by five percentage points reduced hospitalizations by 6.6% and deaths by 6.7%, whereas increasing VC or VE by five percentage points reduced hospitalizations by 3.1% and deaths by 3.2%.

**Conclusions:** Increasing pediatric influenza vaccination coverage back to 2019/20 levels was estimated to significantly reduce pediatric influenza-associated morbidity and mortality.

**What’s known on this subject:** Pediatric influenza vaccination coverage has declined in the United States since the 2019/20 influenza season despite its effectiveness and strong safety record. The impact of this decline on influenza-associated morbidity and mortality has not been quantified.

**What this study adds:** We estimated pediatric influenza-associated outcomes in the 2022/23–2024/25 influenza seasons under different scenarios that increased vaccination coverage and/or effectiveness. We estimated the greatest reductions in influenza-associated hospitalizations and deaths (7.2% and 7.9%, respectively) if coverage remained at 2019/20 levels.

## Introduction

Seasonal influenza causes substantial morbidity and mortality among children <18 years with 12,000– 50,000 hospitalizations and 130–1,160 deaths estimated to occur annually in the United States^1^. The 2024/25 influenza season was particularly impactful and considered high severity in this age group. Cumulative rates of reported pediatric influenza-associated hospitalizations were greater than in any other season since at least 2010/11^2^. In addition, a record number of influenza-associated pediatric deaths were reported since becoming nationally notifiable in 2004^3^. Reported hospitalizations and deaths only include those with laboratory-confirmed influenza and are therefore unlikely to capture the true pediatric burden of severe influenza^4,5^.

Influenza vaccination has a strong safety record and is effective in reducing influenza-associated morbidity and mortality among children^6–11^. Vaccine protection at the population level can be increased through increasing vaccination coverage (VC) or vaccine effectiveness (VE). Influenza VE provides a metric for how well vaccination protects against influenza-associated illness, and varies by season, age group, and disease outcome (for example, protection against mild illness vs. hospitalization). VC reflects the proportion of the population receiving an influenza vaccine in any given season. The highest levels of VC among pediatric age groups in the United States were achieved in the 2019/20 season, with 76% among children 6 months–4 years (surpassing the Healthy People 2030 target of 70%^12^) and 60% among children 5–17 years. However, VC has declined substantially since then, with just 58% of children 6 months–4 years and 48% of children 5–17 years receiving at least one dose of influenza vaccine in the 2024/25 season^13,14^. Of the reported influenza-associated pediatric deaths in 2024/25 who had been eligible for vaccination, approximately 90% were not fully vaccinated against influenza^3^.

We estimate how increased influenza vaccine protection could have impacted pediatric influenza-associated burden in three recent influenza seasons that coincided with declines in VC (2022/23 through 2024/25). We use a previously published compartmental framework to simulate different scenarios for increased VC and/or VE and estimate the reduction in influenza-associated outcomes that could have occurred. Our work explores the potential benefits of influenza vaccination in reducing morbidity and mortality among children.

## Methods

### Influenza-associated burden under observed VC and VE

We estimated the US pediatric influenza-associated burden that occurred in the 2022/23–2024/25 influenza seasons (henceforth referred to as the ‘baseline’ scenario) using a previously published multiplier model^15,16^. We first obtained monthly numbers of laboratory-confirmed influenza-associated hospitalizations for 2022/23–2024/25 from the population-based Influenza Hospitalization Surveillance Network (FluSurv-NET; see Supplementary Information for further details)^17^. These were aggregated into two age groups, 6 months–4 years and 5–17 years, and adjusted for under-detection using information on testing practices and diagnostic test sensitivity^18^. The adjusted influenza-associated hospitalizations were converted to rates by dividing by the total FluSurv-NET catchment population for each age group and season, then translated to total monthly influenza-associated hospitalizations by multiplying by the US population size for each group^19,20^. We estimated the total monthly number of influenza-associated deaths by multiplying the estimated number of hospitalizations by age- and season-specific death-hospitalization ratios that account for under-detection of influenza-associated mortality (Table S1)^1,16^. Finally, we estimated the number of symptomatic illnesses by multiplying the number of hospitalizations by age-specific case-hospitalization ratios, and the number of medically attended illnesses by multiplying symptomatic illnesses by the proportion likely to seek care (Table S1)^1,15,21^. These estimates of illnesses, hospitalizations, and deaths in the baseline scenario represent outcomes that occurred in the presence of influenza vaccine use in the population.

### Impact of influenza vaccination under observed VC and VE

We obtained monthly, age-specific influenza VC estimates for the 2019/20 and 2022/23–2024/25 influenza seasons from FluVaxView^14^. We also obtained age-specific VE estimates for 2022/23–2024/25 from the US Flu Vaccine Effectiveness Network (Flu VE Network), the New Vaccine Surveillance Network (NVSN), and the VIrtual SARS-CoV-2, Influenza, and Other respiratory viruses Network (VISION)^13,22–24^. Further details about these platforms are provided in the Supplementary Information. We distinguished between VE estimated in inpatient vs. outpatient settings and calculated pooled estimates for each (Supplementary Information; Table S2). We used the pooled estimates for VE against outpatient illness when estimating vaccine-mediated protection against symptomatic or medically attended illness, and the pooled estimates for VE against inpatient illness when estimating protection against hospitalization or death. We assumed the vaccine only protected those who were vaccinated (i.e., only had direct effects). We also assumed that vaccine-mediated protection was all-or-nothing, so that the proportion ‘effectively vaccinated’ (i.e., protected) against symptomatic or medically attended illness in each age group and month was the proportion vaccinated in that month multiplied by the corresponding VE against outpatient illness. Similarly, the proportion effectively vaccinated and protected against hospitalization or death was the proportion vaccinated multiplied by the corresponding VE against inpatient illness. We did not account for potential indirect effects, such as a reduction in transmission from vaccinees with influenza virus infection.

We adapted a previously published compartmental framework to estimate the influenza burden prevented by vaccination under observed levels of VC and VE (i.e., the number of outcomes prevented in the baseline scenario) (Figure S1)^25–27^. This framework uses the estimates of illnesses, hospitalizations, and deaths described above, in addition to the VC and VE information. Briefly, for each age group, season, and month, we calculated the number effectively vaccinated against inpatient or outpatient influenza illness as the population who had not yet become symptomatically ill that season multiplied by the respective proportion effectively vaccinated. The population susceptible to hospitalization or death was the number who had not yet become ill or been effectively vaccinated against inpatient illness, and the population susceptible to symptomatic or medically attended illness was the number who had not yet become ill or been effectively vaccinated against outpatient illness. Then, we calculated the risk of each outcome (symptomatic illness, medically attended illness, hospitalization, or death) as the number of outcomes estimated in that month (described above) divided by the corresponding susceptible population from the previous month. Finally, we estimated the number of outcomes that would have occurred in the absence of vaccination using an iterative process in which the monthly risk of each outcome was multiplied by a new susceptible population in which no-one was effectively vaccinated. The number of each outcome prevented by vaccination was then the difference between the number estimated to occur in the presence of vaccination and the number estimated in the absence of vaccination, summed over all months in the season. Further details, including model equations, are provided in the Supplementary Information.

### Impact of influenza vaccination under different VC and/or VE scenarios

We defined four different scenarios to investigate the impact of increased vaccine-mediated protection, assuming this could be achieved by increasing VC, VE, or both. For each season and age group we (1) scaled monthly VC so that final coverage increased by five percentage points; (2) increased VE against outpatient and inpatient illness by five percentage points; (3) increased both VC and final VE by five percentage points (i.e., a combination of scenarios (1) and (2)); and (4) scaled monthly VC so that final coverage increased to that achieved in the 2019/20 season.

Next, we estimated the number of influenza-associated outcomes in each of the four scenarios following previously described methodology^28^. First, we recalculated the proportion effectively vaccinated against inpatient and outpatient illness using the new VC and/or VE inputs defined in each scenario. Then, we iteratively calculated the number of new outcomes as the monthly risk of each outcome (fixed from above) multiplied by a new susceptible population (against inpatient or outpatient illness) that accounted for these re-calculated effectively vaccinated populations. The number of outcomes prevented was then the difference between the new outcomes estimated in each scenario and the outcomes estimated in the baseline scenario, summed over all months in the season.

We generated uncertainty estimates around our point estimates using Monte Carlo simulations. For select model inputs, we drew 10,000 samples from data-informed probability distributions (Table S3) and re-simulated the model for each new set of inputs. Uncertainty is reported as the 95^th^ percentiles of the resulting distributions (referred to as 95^th^ uncertainty intervals [UIs]).

Finally, we described the impact of each scenario using three different metrics: the number of outcomes prevented; the number of outcomes prevented as a percentage of the number of outcomes at baseline; and the number needed to vaccinate (NNV) to prevent one outcome. The latter was calculated as the total number of outcomes prevented in each scenario (i.e., the number of *new* outcomes prevented plus the number of outcomes prevented at baseline) divided by the total number of people vaccinated (whether or not vaccination resulted in effective protection). We note that when a vaccine has only direct vaccine effects, NNV does not change with increasing VC and so we only present NNV estimates for Scenario 2.

All data and model inputs were the result of secondary analyses and considered final for this study as of 23 June 2026 (although the underlying source data may be subject to change with future updates). Analyses were conducted in R version 4.2.2^29^ and unless stated otherwise, absolute numbers are reported to three significant figures and percentages are reported to one decimal place. Results for hospitalizations and deaths are reported in the main text; additional results for medically attended illnesses and symptomatic illnesses are included in the Supplementary Information.

## Results

### Model inputs

Adjusted influenza-associated hospitalization rates, VC, and VE varied by season and age group. Hospitalization rates increased earliest in 2022/23 but reached highest cumulative levels in 2024/25 (Figure S2A). In all seasons, cumulative rates were higher among children 6 months–4 years than children 5–17 years. VC declined each season from 2022/23 to 2024/25 and was always lower among children 5–17 years than children 6 months–4 years (Figure S2B). Although children 5–17 years experienced their lowest VE (against both inpatient and outpatient illness) in 2024/25, children 6 months–4 years experienced their lowest VE in 2022/23 (Figure S2C).

The different scenarios for VC reflected notable increases relative to observed VC (Figure 1, Figure S3). Increasing VC by five percentage points equated to relative increases in VC of between 7.7– 10.5% for any given season and age group, whereas increasing VC to 2019/20 levels reflected relative increases of between 10.0–32.9%. For both scenarios and age groups, the greatest relative increases were realized in the 2024/25 season. Increasing VE against inpatient illness by five percentage points equated to relative increases in VE of between 7.0–11.5% (Figure 2). Similar patterns were observed for VE against outpatient illness, with relative increases between 7.7–11.3% (Figure S4).

**Figure 1.**
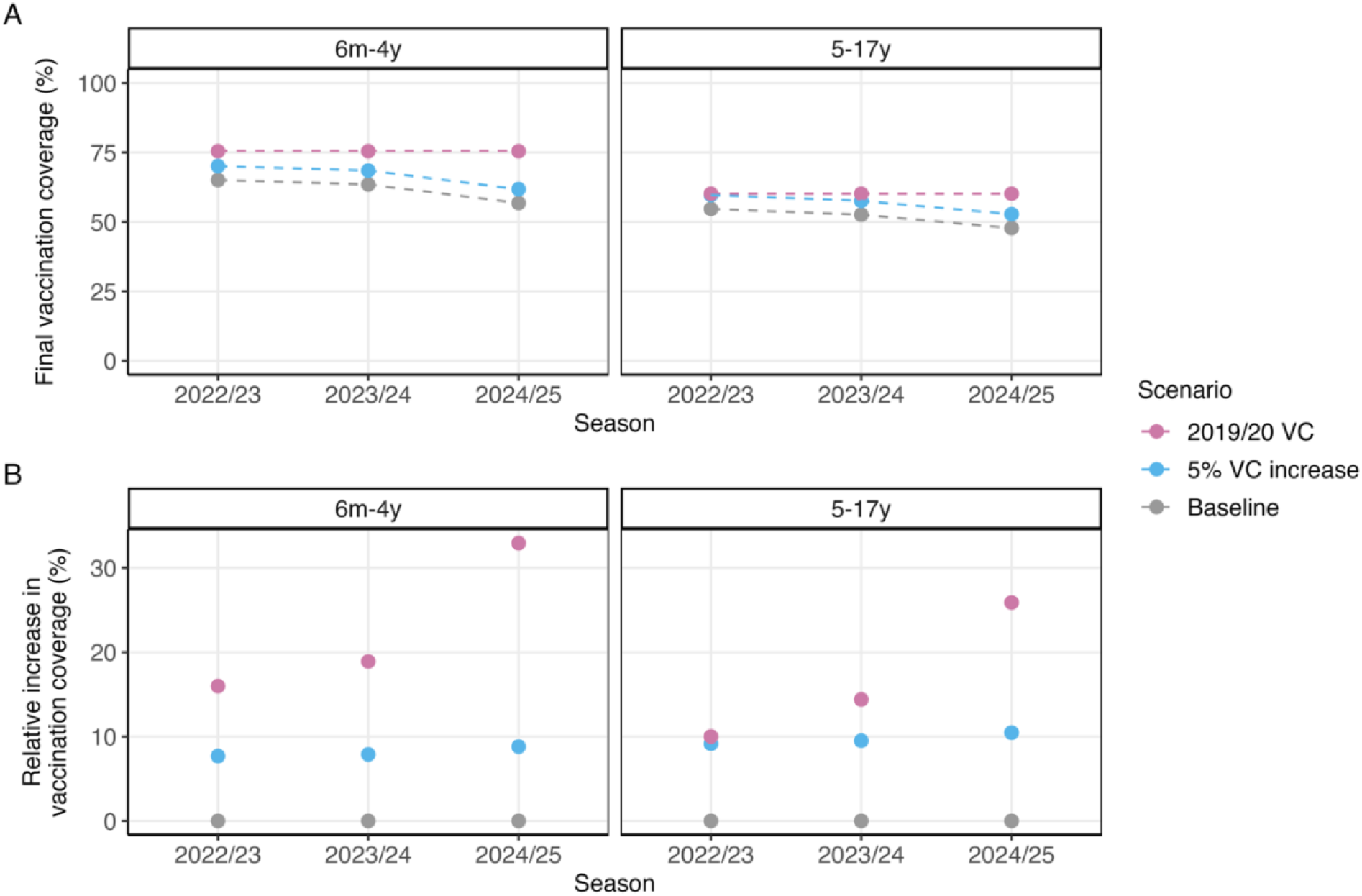
Input values for VC. (A) Values when final VC is increased by five percentage points or to coverage levels achieved in 2019/20. Observed VC (baseline scenario) is shown for comparison. (B) Same as (A) but expressed as the increase relative to baseline. Abbreviations: VC, vaccination coverage.

**Figure 2.**
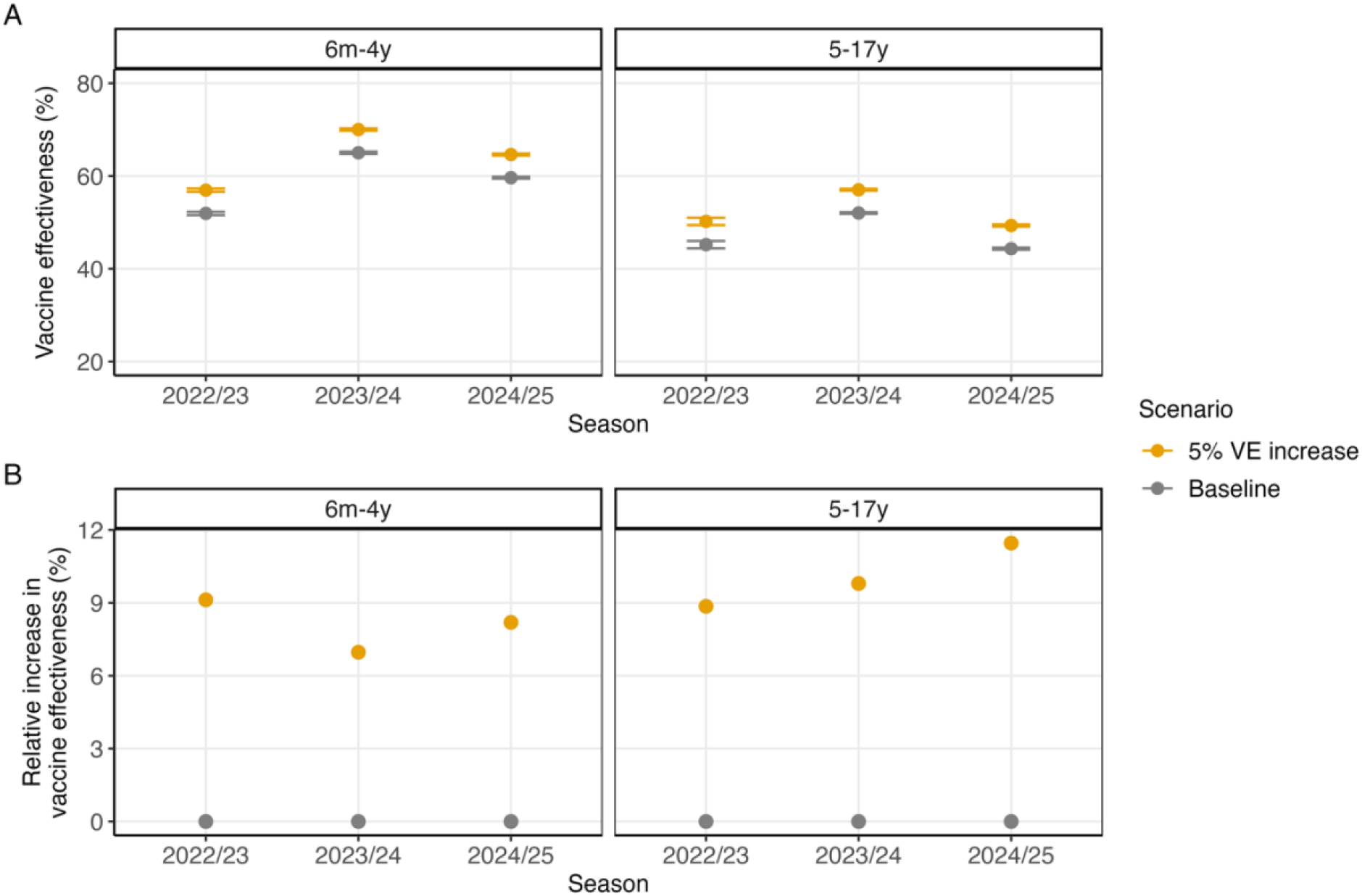
Input values for VE against inpatient illness. (A) Values when VE against inpatient illness is increased by five percentage points. Observed VE (baseline scenario) is shown for comparison. Error bars represent 95% confidence intervals. (B) Same as (A) but expressed as the increase relative to baseline. Abbreviations: VE, vaccine effectiveness.

### Estimated influenza-associated outcomes and impact of different vaccination scenarios

Among all children 6 months–17 years, we estimated there were 120,000 (95^th^ UI: 116,000–124,000) influenza-associated hospitalizations and 1,650 (1,160–2,520) deaths across the three seasons (Figure S5). The greatest single-season burden was estimated in 2024/25, with 46,600 (44,300–49,100) hospitalizations and 725 (401–1,360) deaths. Increasing VC by five percentage points (Scenario 1) resulted in 3,770 fewer estimated hospitalizations (95^th^ UI: 3,590–3,950) and 54 (36–78) fewer deaths across all seasons (3.1% and 3.2% reductions relative to baseline, respectively) (Figures 3 and 4; see also Figures S6 and S7, and Tables S4–S7). Increasing VE by five percentage points (Scenario 2) had similar estimated impacts to Scenario 1 across the three seasons. In Scenario 2, the NNV to prevent one hospitalization decreased from 2,700 at baseline to 2,480, and the NNV to prevent one death decreased from 190,000 to 174,000 (Figures S8 and S9; see also Figures S10 and S11).

**Figure 3.**
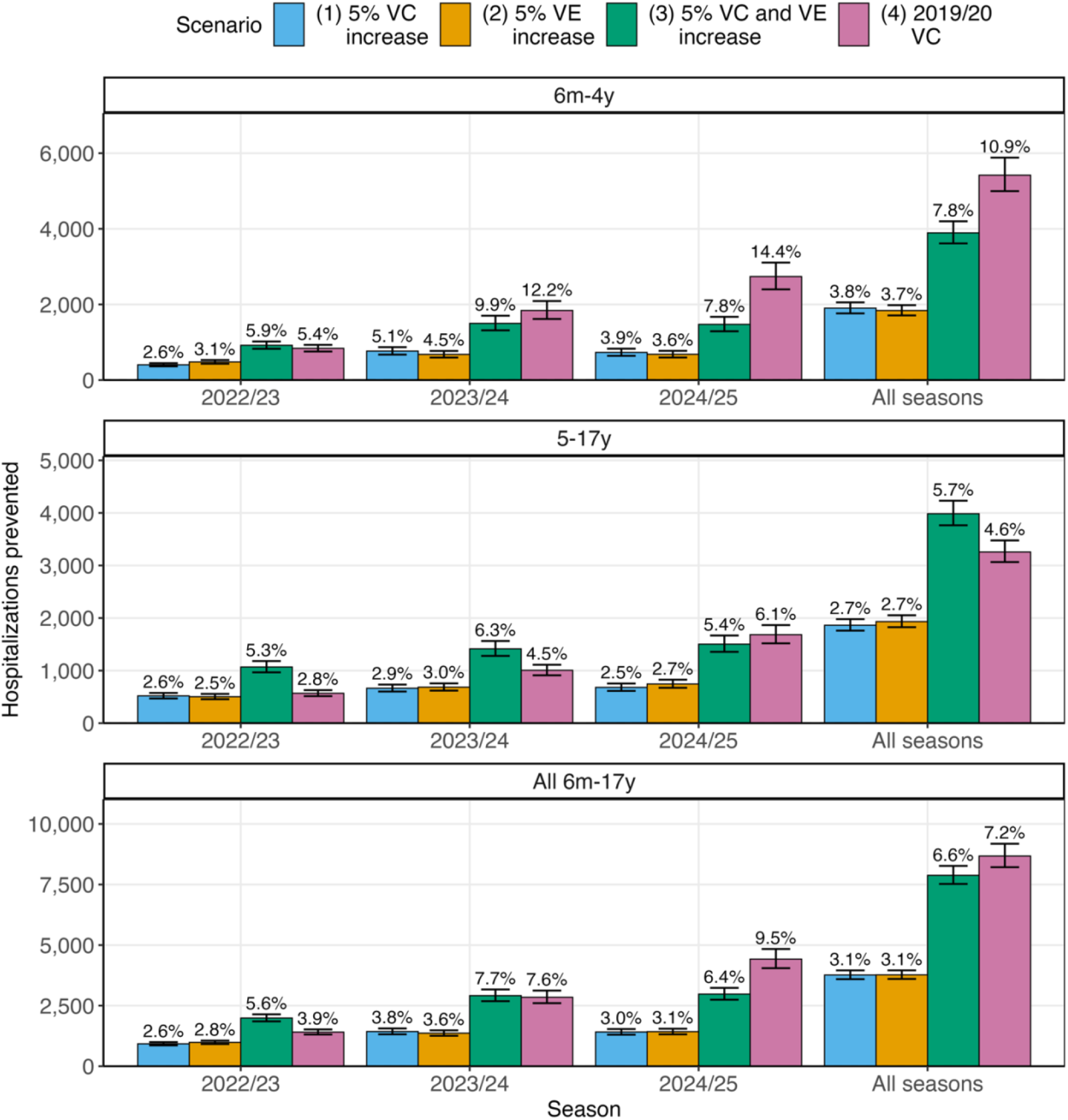
Estimated influenza-associated hospitalizations prevented by vaccination in different vaccination scenarios. Bars represent point estimates and error bars reflect 95^th^ uncertainty intervals. Numbers above bars are point estimates of the percentage reduction in hospitalizations relative to baseline. Abbreviations: VC, vaccination coverage; VE, vaccine effectiveness.

**Figure 4.**
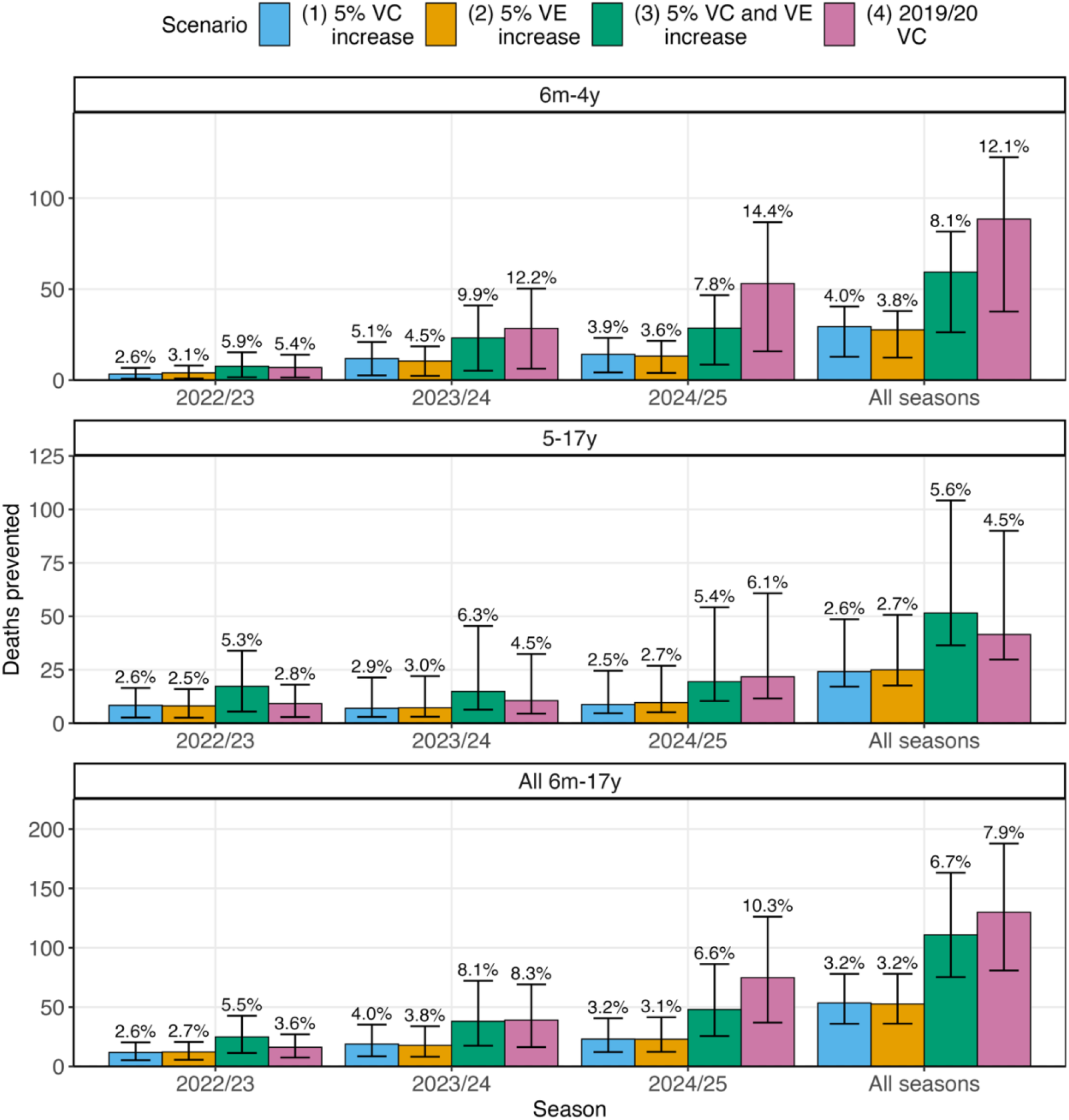
Estimated influenza-associated deaths prevented by vaccination in different vaccination scenarios. Bars represent point estimates and error bars reflect 95^th^ uncertainty intervals. Numbers above bars are point estimates of the percentage reduction in deaths relative to baseline. Abbreviations: VC, vaccination coverage; VE, vaccine effectiveness.

Increasing both VC and VE by five percentage points (Scenario 3) prevented more outcomes than increasing either in isolation (Figures 3 and 4; Tables S4–S7). The greatest single-season impacts were estimated in 2024/25. Across all seasons, an estimated 7,880 (95^th^ UI: 7,520–8,270) fewer hospitalizations and 111 (75–163) fewer deaths would have occurred (reductions of 6.6% and 6.7%, respectively). Increasing VC to 2019/20 levels (Scenario 4) also had a substantially greater impact than Scenarios 1 or 2 for most seasons (Figures 3 and 4). Similar to Scenario 3, the greatest reductions were estimated in 2024/25. Across all seasons, 8,670 (95^th^ UI: 8,210–9,180) fewer hospitalizations and 130 (81–188) fewer deaths were estimated to have occurred (7.2% and 7.9% reductions, respectively). This was the greatest estimated impact of the scenarios considered.

## Discussion

We aimed to estimate how recent decreases in influenza vaccine coverage among children could have impacted US pediatric influenza burden in the 2022/23–2024/25 influenza seasons. By combining a compartmental framework with different scenarios for VC and VE, we explored the relative contributions of increasing VC or VE on reducing influenza burden among children. Increasing VC or VE by five percentage points reduced the number of influenza-associated hospitalizations by 3.1% and deaths by 3.2% across all seasons and age groups, and increasing both VC and VE by five percentage points reduced these outcomes by 6.6 and 6.7%, respectively. Setting VC to levels achieved in 2019/20, prior to recent declines, had the greatest impact, reducing hospitalizations by 7.2% and deaths by 7.9% (equivalent to 8,670 pediatric hospitalizations and 130 pediatric deaths across all three seasons). These results suggest that increasing influenza vaccination protection in pediatric populations could have substantially decreased influenza-associated morbidity and mortality in the 2022/23–2024/25 seasons and highlights the potential impact of improving both vaccine coverage and vaccine effectiveness in future seasons.

Seasonal influenza causes considerable disease burden in children. At observed levels of VC and VE, we estimated there were a total of 120,000 influenza-associated pediatric hospitalizations and 1,650 deaths from 2022/23–2024/25. In particular, the 2024/25 season was classified as high severity in pediatric populations and had a record number of influenza-associated pediatric deaths reported, in addition to unexplained increases in pediatric influenza-associated encephalopathy^30^. The 2025/26 season has also been classified as high severity in children, suggesting another season with substantial pediatric disease burden^31^. While children younger than 5 years old (especially those younger than 2), and children of any age with certain chronic health conditions, are at increased risk of severe influenza-associated outcomes^32,33^, it is important to note that all children are at risk. For example, of the pediatric deaths reported in 2024/25 among children with known medical history, 43% occurred in those without any medical conditions associated with higher risk of influenza complications^34^. Similarly, 46% of children hospitalized for influenza in 2024/25 had no known conditions associated with higher risk^35^.

Influenza vaccination is an important way to protect children against influenza-associated morbidity and mortality, with strong evidence supporting its safety and effectiveness^6–11,36,37^. Notably, of the reported pediatric influenza-associated deaths in 2024/25 who had been eligible for vaccination, approximately 90% were not fully vaccinated against influenza^3^. The highest levels of VC among children were observed in 2019/20. However, VC declined in each subsequent season to 2024/25, and preliminary coverage estimates indicate that it remained low in 2025/26^38^. Our results suggest that increasing pediatric VC to levels previously achieved during the 2019/20 influenza season could have substantially reduced influenza-associated morbidity and mortality and may help inform future discussions between healthcare providers and families about the decision to vaccinate children against influenza.

Our scenarios for increased VC and/or VE were designed to reflect attainable goals. The scenario assuming 2019/20 VC is based on coverage achieved in a recent prior season, and our scenario assuming a five percentage point VC increase is small compared with the 18 and 12 percentage-point decreases observed from 2019/20 to 2024/25 among children 6 months–4 years and 5–17 years, respectively. With respect to VE, a five percentage point increase is within the range of typical inter-seasonal variation^39^. Consistent increases in VE may be challenging to achieve in the short term, in part due to virus evolution and potential antigenic differences between circulating and vaccine strains. However, progress could be achieved through the development of more immunogenic influenza vaccines. Further studies could also assess the impact of increasing use of certain currently licensed influenza vaccines, such as live attenuated or cell-based influenza vaccines that are potentially more effective than standard egg-based vaccines in children^40–42^. That being said, we still found substantial reductions in morbidity and mortality when VE remained at observed levels and only VC was increased. Our scenario that combined 5% increases in VC and VE highlights how the impact of increased coverage could be further enhanced in seasons with high VE.

Limitations of the original framework to estimate vaccine-prevented influenza-associated burden have been discussed previously^25,27^. First, the case-hospitalization ratios and medically attended fractions were estimated during the 2009 pandemic and may have changed due to underlying variation in disease severity and care seeking behaviors^43,44^. Although such differences could impact our estimates of the number of symptomatic and medically attended illnesses prevented, estimates of the relative reductions in these outcomes should remain unchanged. Second, FluSurv-NET covers approximately 10% of the US population. While the catchment area is demographically similar to the US population, the data may not be representative of the country as a whole. Third, the VC data were based on parent-reported vaccination status and may be influenced by recall bias^45,46^. In addition, these data represent the proportion of children receiving at least one dose of seasonal influenza vaccine, although two doses are recommended for children 8 years and younger receiving their first seasonal influenza vaccine, for children who have only ever received one dose in previous seasons, or for children with unknown vaccination history^36^. We may have overestimated the number effectively vaccinated if these children require two doses to receive complete protection from vaccination. Fourth, our framework is conservative in that it does not consider any indirect benefits that would have been gained if vaccination reduced the risk of onward transmission from infected vaccinated individuals. Finally, we acknowledge that underlying data for the model inputs may be subject to change. In particular, data for the 2023/24 and 2024/25 seasons were considered preliminary as of 23 June 2026. While our estimates of the absolute numbers of outcomes prevented may be sensitive to changes in these data, the relative reductions in outcomes should be less so.

## Conclusion

Influenza VC among pediatric age groups declined substantially from 2019/20 to 2024/25. The 2024/25 influenza season was also particularly severe, with the highest number of influenza-associated pediatric deaths ever reported. Here we estimated that increasing influenza vaccination coverage among children back to 2019/20 levels could have substantially reduced influenza-associated morbidity and mortality in the 2022/23–2024/25 seasons. Our findings support that seasonal vaccination is an important way to protect children against influenza-associated morbidity and mortality.

## Data Availability

All data were the result of secondary analyses and used solely as model inputs. Data available online are cited within the text, and available at https://www.cdc.gov/flu-burden/php/data-vis-vac/index.html and https://www.cdc.gov/flu-burden/php/data-vis/index.html.

https://www.cdc.gov/flu-burden/php/data-vis/index.html

