## Supplementary Information for "Potential impact of changes in influenza vaccination coverage and effectiveness on pediatric influenza burden in the United States, 2022/23–2024/25"

#### Surveillance Platforms

*Influenza Hospitalization Surveillance Network (FluSurv-NET)*

FluSurv-NET conducts surveillance on laboratory-confirmed influenza-associated hospitalizations through a network of hospitals in fourteen states. The network represents approximately 34 million people (or 10% of the US population). Cases are defined as patients living in a FluSurv-NET catchment area who receive a positive laboratory-confirmed influenza test within 14 days prior to, or during, hospitalization. FluSurv-NET activity was reviewed by the US Centers for Disease Control and Prevention (CDC), deemed not research, and was conducted consistent with applicable federal law and CDC policy: 45 C.F.R. part 46.102(l)(2), 21 C.F.R. part 56; 42 U.S.C. Sect. 241(d); 5 U.S.C. Sect. 552a; 44 U.S.C. Sect. 3501 et seq. FluSurv-NET sites obtained human subjects and ethics approval from their respective state health department, academic partner, and participating hospital institutional review boards as needed. Further information can be found at: https://www.cdc.gov/fluview/overview/influenza-hospitalization-surveillance.html.

*US Flu Vaccine Effectiveness Network (Flu VE Network)*

The Flu VE Network conducts surveillance on influenza-associated outpatient illness through a network of seven sites in seven states (and one coordinating center) and produces estimates of VE against outpatient illness. The study protocol was reviewed by CDC, determined to be public health surveillance, and conducted consistent with applicable federal law and CDC policy (45 CFR 46.102[l][2]). Further information can be found at: https://www.cdc.gov/flu-vaccines-work/php/vaccine-effectiveness/us-flu-ve-network.html.

*New Vaccine Surveillance Network (NVSN)*

NVSN conducts surveillance on acute respiratory illness (including influenza-associated inpatient and outpatient illness) through a network of seven pediatric health systems in seven states. NVSN produces estimates of VE against outpatient and inpatient illness. The study protocol was reviewed and approved by the CDC and the site institutional review boards. This activity was reviewed by CDC, deemed not research, and was conducted consistent with applicable federal law and CDC policy (see e.g., 45 CFR part 46.102(l)(2), 21 CFR part 56; 42 USC Sect. 241(d); 5 USC Sect. 552a; 44 USC Sect. 3501 et seq). Further information can be found at: https://www.cdc.gov/nvsn/php/about/index.html.

*Virtual SARS-CoV-2, Influenza, and Other respiratory viruses Network (VISION)*

VISION conducts surveillance on influenza-associated inpatient and outpatient outcomes through a network of eight sites in nine states that cover a variety of healthcare settings including urgent care, emergency departments and hospitals. VISION produces estimates of VE against outpatient illness and inpatient illness. The VISION activity was reviewed by CDC, deemed not research or research not involving human subjects, and was conducted consistent with applicable federal law and CDC policy (See e.g., 45C.F.R. part 46, 21C.F.R. part 56; 42 U.S.C. §241(d); 5 U.S.C. §552a; 44 U.S.C. §3501 et seq). Further information can be found at: https://www.cdc.gov/flu-vaccines-work/php/vaccine-effectiveness/vision-network.html.

**Pooled VE estimates**

For each platform, setting (inpatient or outpatient), age group, and season, we assumed VE followed a normal distribution with mean and standard deviation equal to the corresponding mean and standard error of the original VE estimate, respectively. We then simulated 500 samples from these distributions and calculated pooled VE estimates as the median VE for each setting, age group and season; sampled distributions from different platforms were combined if they had age group, season, and setting in common.

**Model**

*With observed vaccination*

For each age group, season, and month, m, we first calculated the number of people effectively vaccinated against outpatient illness ($EV_{out}$) as

$$EV_{out,m} = \left(P - \sum_{j=1}^{m} C_{j-1}\right) \times VC_m \times VE_{out},$$

where P is the population size (assuming everyone is susceptible at the start of the season), $\sum_{j=1}^{m} C_{j-1}$ is the cumulative number of estimated symptomatic illnesses up to the prior month (with $C_0 = 0$), $VC_m$ is the proportion vaccinated in the current month, and $VE_{out}$ is the VE against outpatient illness. Similarly, the number of people effectively vaccinated against inpatient illness ($EV_{in}$) is

$$EV_{in,m} = \left(P - \sum_{j=1}^{m} C_{j-1}\right) \times VC_m \times VE_{in},$$

where $VE_{in}$ is the VE against inpatient illness.

The number susceptible ($S_{out}$) against outpatient illness is estimated as

$$S_{out,m} = P - \sum_{j=1}^{m} C_j - \sum_{j=1}^{m} EV_{out,j},$$

and the number susceptible against inpatient illness ($S_{in}$) as

$$S_{in,m} = P - \sum_{j=1}^{m} C_j - \sum_{j=1}^{m} EV_{in,j},$$

where $S_{out,0} = S_{in,0} = P$. We then calculated the risk of symptomatic illness ($R_C$) and medically attended illness ($R_{MA}$) using $S_{out}$, and the risk of hospitalization ($R_H$) and death ($R_D$) using $S_{in}$. The expressions are as follows:

$$R_{C,m} = \frac{C_m}{S_{out,m-1}},$$

$$R_{MA,m} = \frac{MA_m}{S_{out,m-1}},$$

$$R_{H,m} = \frac{H_m}{S_{in,m-1}},$$

$$R_{D,m} = \frac{D_m}{S_{in,m-1}},$$

where MA, H, D are the estimated number of medically attended illnesses, hospitalizations, and deaths, respectively.

*Without vaccination*

The risks defined above are fixed and used to calculate the number of each outcome in the absence of vaccination. For example, the new number of symptomatic illnesses is given by

$$\widehat{C_m} = \widehat{S_m} \times R_{C,m},$$

where $\widehat{S_m}$ is the susceptible population without vaccination, so that

$$\widehat{S_m} = \widehat{S_{m-1}} - \widehat{C_{m-1}},$$

with $\widehat{S_0}$= P and $\widehat{C_0}$= 0. Note that there is now only one susceptible population for all outcomes since there is no effectively vaccinated population that requires distinguishing between those susceptible to inpatient vs. outpatient outcomes.

We obtained similar expressions for the number of new medically attended illnesses, hospitalizations, and deaths, i.e.,

$$\widehat{MA_m} = \widehat{S_m} \times R_{MA,m},$$

$$\widehat{H_m} = \widehat{S_m} \times R_{H,m},$$

and

$$\widehat{D_m} = \widehat{S_m} \times R_{D,m},$$

respectively. Finally, the number of each outcome prevented by vaccination is the number of outcomes in the absence of vaccination minus the number estimated in the presence of vaccination. For example, $\widehat{C_m} - C_m$. The total number of outcomes prevented is then summed across all months, for each age group and season.

### Supplementary Tables

**Table S1. Point value inputs for the influenza case-hospitalization ratio (CHR), death-hospitalization ratio (DHR), and fraction of symptomatic illnesses that are medically attended (MA fraction).** The CHR and MA fraction are fixed across all seasons.

| Age group | Season | CHR | DHR | MA fraction |
|---|---|---|---|---|
| 6 months–4 years | 2022/23 | 143.4 | 0.008 | 0.67 |
| | 2023/24 | | 0.015 | |
| | 2024/25 | | 0.019 | |
| 5–17 years | 2022/23 | 364.7 | 0.016 | 0.52 |
| | 2023/24 | | 0.011 | |
| | 2024/25 | | 0.013 | |

**Table S2. Influenza vaccine effectiveness surveillance platforms and settings for which estimates are produced.** Outpatient settings include emergency departments, urgent care centers, and clinics.

| Surveillance platform | Setting for VE estimate |
|---|---|
| Flu VE Network | Outpatient |
| NVSN | Inpatient and outpatient |
| VISION | Inpatient and outpatient |

**Table S3. Parameter distributions for Monte Carlo simulations.** Abbreviations: $l_{95}$, lower 95th percentile; $u_{95}$, upper 95th percentile; SE, standard error; VC, vaccine coverage; VE, vaccine effectiveness.

| Input | Stratifications | Distribution | Parameter |
|---|---|---|---|
| Number of FluSurv-NET hospitalizations | Season, month, age | Poisson | $\lambda$ = observed number |
| Outcome multipliers | Season, age | | |
| Under-detection | | Scaled Beta distribution | Range = [observed $l_{95}$, observed $u_{95}$] |
| Medically attended fraction | | Normal | $\mu$ = observed fraction<br>$\sigma$ = observed SE |
| Death-hospitalization ratio | | See Rolfes et al.[1] | |

[1] Rolfes MA, Foppa IM, Garg S, et al. Annual estimates of the burden of seasonal influenza in the United States: A tool for strengthening influenza surveillance and preparedness. Influenza Other Respir Viruses. 2018;12(1):132-137. doi:10.1111/irv.12486

| | | | |
|---|---|---|---|
| VC | Season, month, age | Normal | $\mu$ = observed fraction<br>$\sigma$ = observed standard error |
| VE* | Season, age | Normal | $\mu$ = observed log(1 – VE)<br>$\sigma$ = observed SE of log(1 – VE) |

*Equivalent expressions for influenza VE against outpatient and inpatient illness.

**Table S4. Estimated influenza-associated hospitalizations prevented by vaccination in different vaccination scenarios.** Point estimates are shown with the percentage reduction relative to baseline in parentheses. Abbreviations: VC, vaccination coverage; VE, vaccine effectiveness.

| Season | Age | Scenario | | | |
|---|---|---|---|---|---|
| | | 5% VC increase | 5% VE increase | 5% VC and VE increase | 2019/20 VC |
| 2022/23 | 6m-4y | 404 (2.6%) | 479 (3.1%) | 920 (5.9%) | 839 (5.4%) |
| 2023/24 | 6m-4y | 767 (5.1%) | 678 (4.5%) | 1,500 (9.9%) | 1,840 (12.%) |
| 2024/25 | 6m-4y | 732 (3.9%) | 681 (3.6%) | 1,470 (7.8%) | 2,740 (14.%) |
| All seasons | 6m-4y | 1,900 (3.8%) | 1,840 (3.7%) | 3,890 (7.8%) | 5,420 (11.%) |
| 2022/23 | 5-17y | 519 (2.6%) | 503 (2.5%) | 1,070 (5.3%) | 568 (2.8%) |
| 2023/24 | 5-17y | 664 (2.9%) | 684 (3.0%) | 1,410 (6.3%) | 1,010 (4.5%) |
| 2024/25 | 5-17y | 680 (2.5%) | 745 (2.7%) | 1,500 (5.4%) | 1,680 (6.1%) |
| All seasons | 5-17y | 1,860 (2.7%) | 1,930 (2.7%) | 3,980 (5.7%) | 3,260 (4.6%) |
| 2022/23 | All 6m-17y | 923 (2.6%) | 982 (2.8%) | 1,990 (5.6%) | 1,410 (3.9%) |
| 2023/24 | All 6m-17y | 1,430 (3.8%) | 1,360 (3.6%) | 2,910 (7.7%) | 2,850 (7.6%) |
| 2024/25 | All 6m-17y | 1,410 (3.0%) | 1,430 (3.1%) | 2,980 (6.4%) | 4,420 (9.5%) |
| All seasons | All 6m-17y | 3,770 (3.1%) | 3,770 (3.1%) | 7,880 (6.6%) | 8,670 (7.2%) |

**Table S5. Estimated influenza-associated deaths prevented by vaccination in different vaccination scenarios.** Point estimates are shown with the percentage reduction relative to baseline in parentheses. Abbreviations: VC, vaccination coverage; VE, vaccine effectiveness.

| Season | Age | Scenario | | | |
|---|---|---|---|---|---|
| | | 5% VC increase | 5% VE increase | 5% VC and VE increase | 2019/20 VC |
| 2022/23 | 6m-4y | 3 (2.6%) | 4 (3.1%) | 8 (5.9%) | 7 (5.4%) |
| 2023/24 | 6m-4y | 12 (5.1%) | 10 (4.5%) | 23 (9.9%) | 28 (12.2%) |
| 2024/25 | 6m-4y | 14 (3.9%) | 13 (3.6%) | 29 (7.8%) | 53 (14.4%) |
| All seasons | 6m-4y | 29 (4.0%) | 28 (3.8%) | 59 (8.1%) | 88 (12.1%) |
| 2022/23 | 5-17y | 8 (2.6%) | 8 (2.5%) | 17 (5.3%) | 9 (2.8%) |
| 2023/24 | 5-17y | 7 (2.9%) | 7 (3.0%) | 15 (6.3%) | 11 (4.5%) |
| 2024/25 | 5-17y | 9 (2.5%) | 10 (2.7%) | 19 (5.4%) | 22 (6.1%) |
| All seasons | 5-17y | 24 (2.6%) | 25 (2.7%) | 52 (5.6%) | 42 (4.5%) |
| 2022/23 | All 6m-17y | 12 (2.6%) | 12 (2.7%) | 25 (5.5%) | 16 (3.6%) |

| | | | | | |
|---|---|---|---|---|---|
| 2023/24 | All 6m-17y | 19 (4.0%) | 18 (3.8%) | 38 (8.1%) | 39 (8.3%) |
| 2024/25 | All 6m-17y | 23 (3.2%) | 23 (3.1%) | 48 (6.6%) | 75 (10.3%) |
| All seasons | All 6m-17y | 54 (3.2%) | 53 (3.2%) | 111 (6.7%) | 130 (7.9%) |

**Table S6. Estimated influenza-associated medically attended illnesses prevented by vaccination in different vaccination scenarios.** Point estimates are shown with the percentage reduction relative to baseline in parentheses. Abbreviations: VC, vaccination coverage; VE, vaccine effectiveness.

| Season | Age | Scenario | | | |
|---|---|---|---|---|---|
| | | 5% VC increase | 5% VE increase | 5% VC and VE increase | 2019/20 VC |
| 2022/23 | 6m-4y | 36,000 (2.4%) | 45,200 (3.0%) | 84,700 (5.7%) | 75,000 (5.0%) |
| 2023/24 | 6m-4y | 62,700 (4.3%) | 61,300 (4.2%) | 129,000 (8.9%) | 151,000 (10.4%) |
| 2024/25 | 6m-4y | 68,000 (3.7%) | 64,800 (3.6%) | 139,000 (7.6%) | 255,000 (13.9%) |
| All seasons | 6m-4y | 167,000 (3.5%) | 171,000 (3.6%) | 352,000 (7.4%) | 480,000 (10.1%) |
| 2022/23 | 5-17y | 74,100 (1.9%) | 89,600 (2.4%) | 172,000 (4.5%) | 81,000 (2.1%) |
| 2023/24 | 5-17y | 129,000 (3.0%) | 131,000 (3.1%) | 272,000 (6.4%) | 196,000 (4.6%) |
| 2024/25 | 5-17y | 132,000 (2.5%) | 142,000 (2.7%) | 288,000 (5.5%) | 325,000 (6.2%) |
| All seasons | 5-17y | 335,000 (2.5%) | 362,000 (2.7%) | 732,000 (5.5%) | 602,000 (4.5%) |
| 2022/23 | All 6m-17y | 110,000 (2.1%) | 135,000 (2.5%) | 257,000 (4.8%) | 156,000 (2.9%) |
| 2023/24 | All 6m-17y | 192,000 (3.3%) | 192,000 (3.3%) | 401,000 (7.0%) | 346,000 (6.0%) |
| 2024/25 | All 6m-17y | 200,000 (2.8%) | 207,000 (2.9%) | 427,000 (6.0%) | 580,000 (8.2%) |
| All seasons | All 6m-17y | 502,000 (2.8%) | 533,000 (2.9%) | 1,080,000 (6.0%) | 1,080,000 (6.0%) |

**Table S7. Estimated influenza-associated symptomatic illnesses prevented by vaccination in different vaccination scenarios.** Point estimates are shown with the percentage reduction relative to baseline in parentheses. Abbreviations: VC, vaccination coverage; VE, vaccine effectiveness.

| Season | Age | Scenario | | | |
|---|---|---|---|---|---|
| | | 5% VC increase | 5% VE increase | 5% VC and VE increase | 2019/20 VC |
| 2022/23 | 6m-4y | 53,800 (2.4%) | 67,400 (3.0%) | 126,000 (5.7%) | 112,000 (5.0%) |
| 2023/24 | 6m-4y | 93,600 (4.3%) | 91,400 (4.2%) | 192,000 (8.9%) | 225,000 (10.4%) |
| 2024/25 | 6m-4y | 102,000 (3.7%) | 96,800 (3.6%) | 207,000 (7.6%) | 380,000 (13.9%) |
| All seasons | 6m-4y | 249,000 (3.5%) | 256,000 (3.6%) | 525,000 (7.4%) | 716,000 (10.1%) |
| 2022/23 | 5-17y | 142,000 (1.9%) | 172,000 (2.4%) | 331,000 (4.5%) | 156,000 (2.1%) |
| 2023/24 | 5-17y | 248,000 (3.0%) | 251,000 (3.1%) | 523,000 (6.4%) | 376,000 (4.6%) |
| 2024/25 | 5-17y | 253,000 (2.5%) | 273,000 (2.7%) | 554,000 (5.5%) | 626,000 (6.2%) |
| All seasons | 5-17y | 644,000 (2.5%) | 696,000 (2.7%) | 1,410,000 (5.5%) | 1,160,000 (4.5%) |
| 2022/23 | All 6m-17y | 196,000 (2.1%) | 240,000 (2.5%) | 457,000 (4.8%) | 268,000 (2.8%) |
| 2023/24 | All 6m-17y | 342,000 (3.3%) | 343,000 (3.3%) | 716,000 (6.9%) | 601,000 (5.8%) |

| | | | | | |
|---|---|---|---|---|---|
| 2024/25 | All 6m-17y | 354,000 (2.8%) | 370,000 (2.9%) | 761,000 (5.9%) | 1,010,000 (7.9%) |
| All seasons | All 6m-17y | 893,000 (2.7%) | 952,000 (2.9%) | 1,930,000 (5.9%) | 1,870,000 (5.7%) |

**Supplementary Figures**

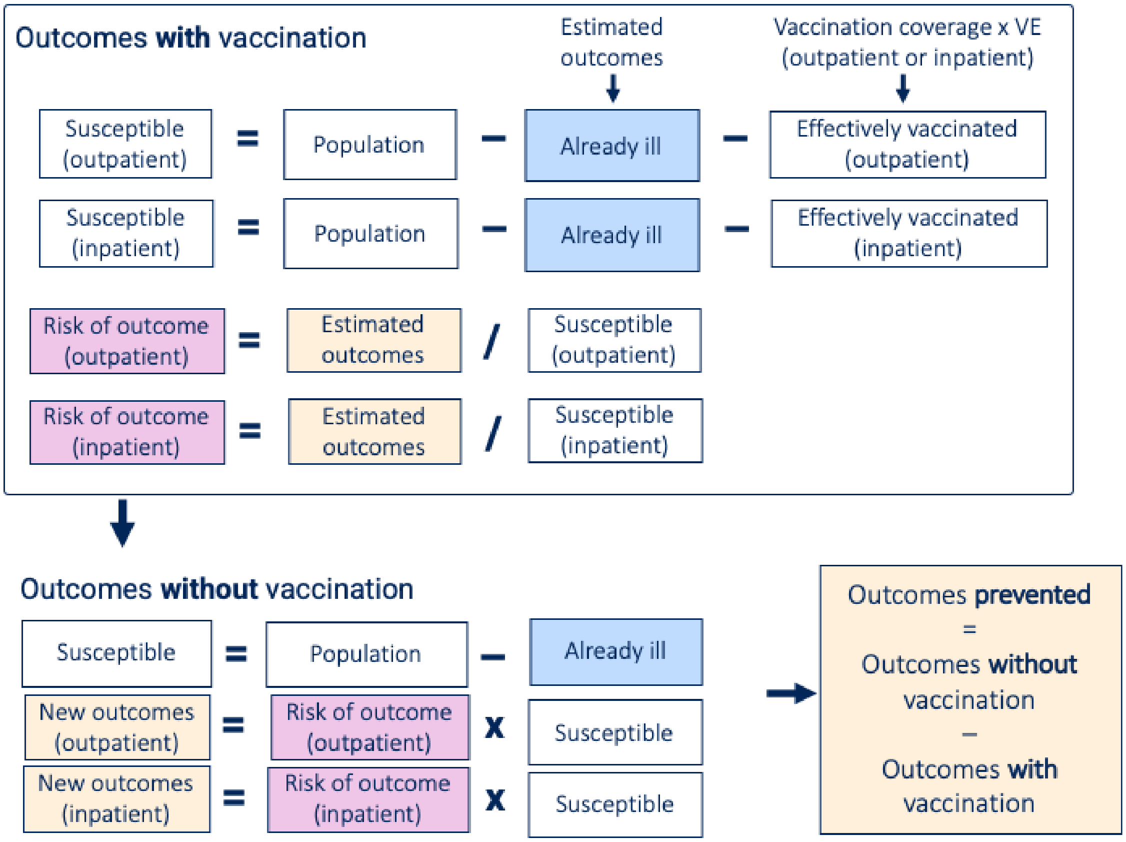


**Figure S1. Schematic of the compartmental framework for estimating influenza vaccine-prevented burden under observed VC and VE.** Abbreviations: VC, vaccination coverage; VE vaccine effectiveness.

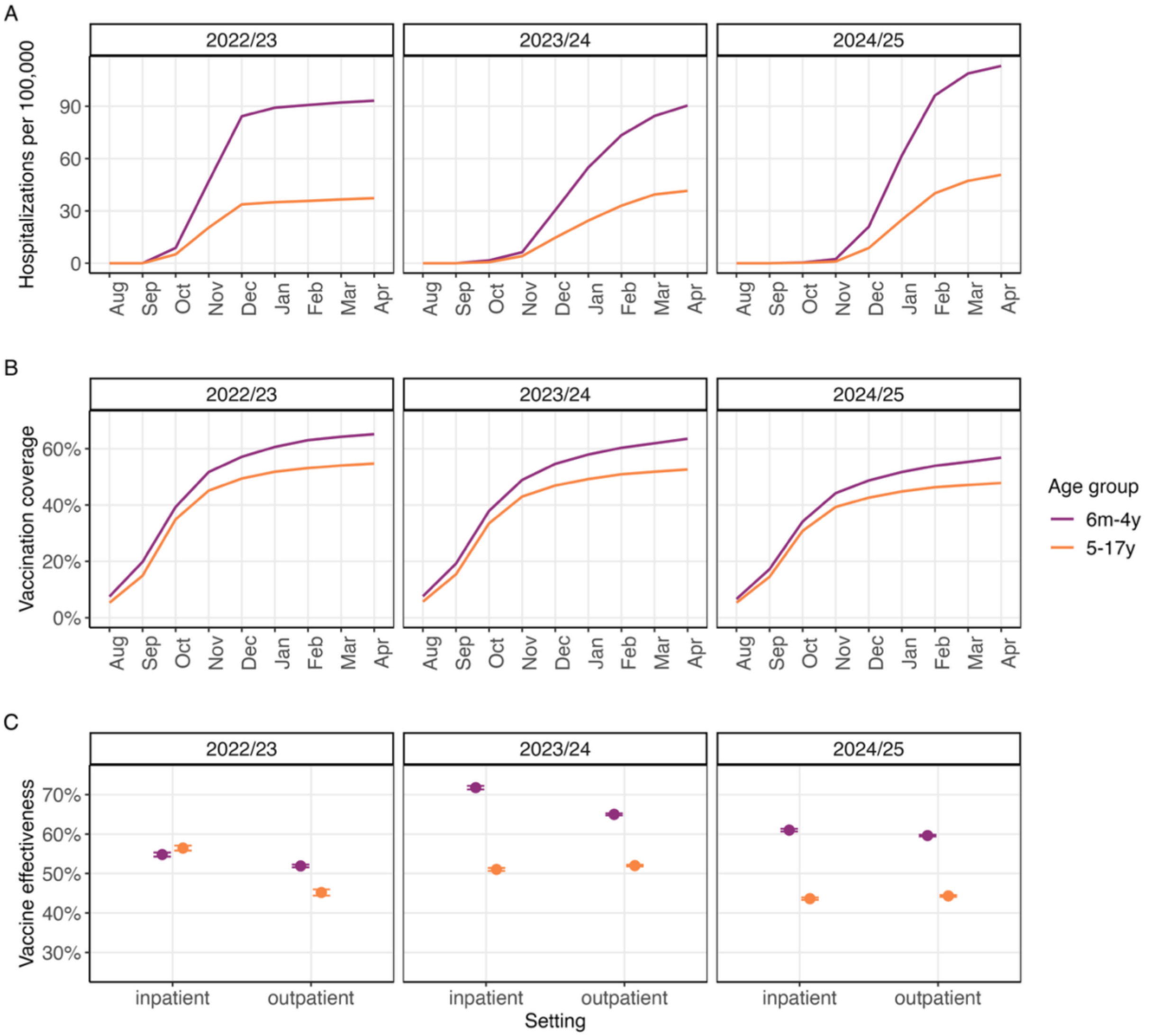


**Figure S2. Adjusted influenza-associated hospitalization rates, vaccination coverage (VC), and vaccine effectiveness (VE) estimates for the 2022/23–2024/25 influenza seasons.** (A) Cumulative rates of laboratory-confirmed influenza-associated hospitalizations per 100,000. Rates are adjusted for under-reporting using information on testing practices and test sensitivity. (B) Cumulative influenza VC. (C) Pooled estimates of influenza VE against inpatient and outpatient illness. Error bars represent 95% confidence intervals.

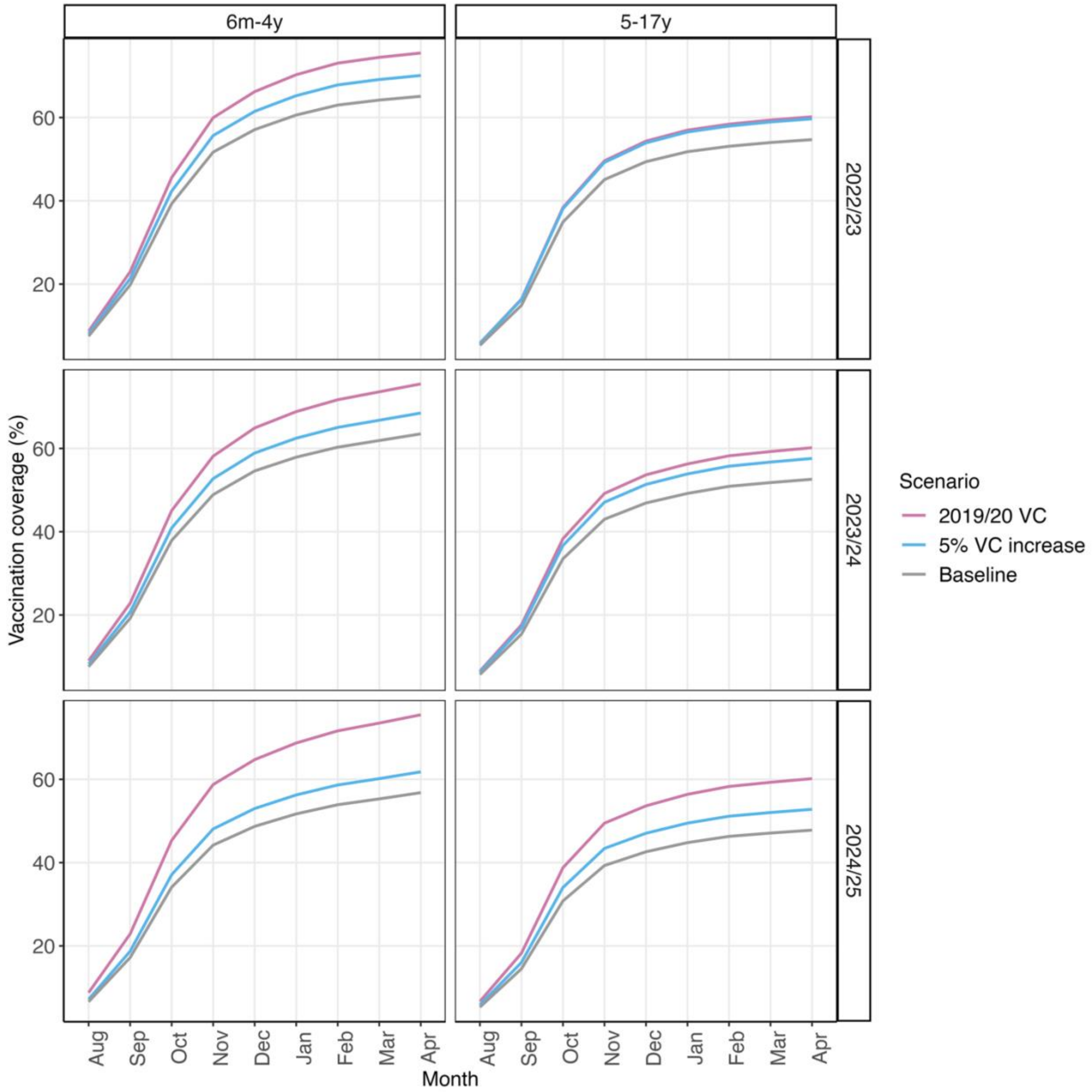


**Figure S3. Input values for increased VC.** Monthly cumulative VC when final VC is increased by five percentage points or to levels achieved in 2019/20. Observed VC (baseline scenario) is shown for comparison. Abbreviations: VC, vaccination coverage.

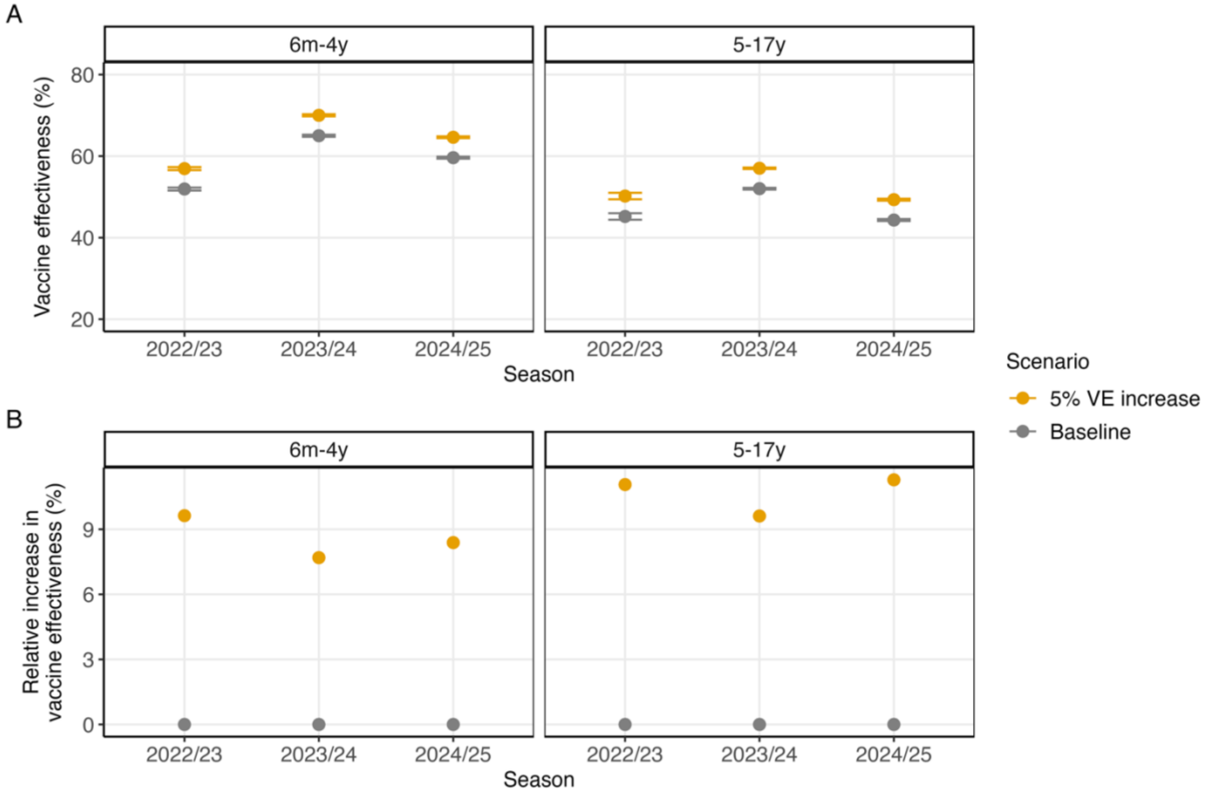


**Figure S4. Input values for VE against outpatient illness.** (A) Values when VE against outpatient illness is increased by five percentage points. Observed VE (baseline scenario) is shown for comparison. Error bars represent 95% confidence intervals. (B) Same as (A) but expressed as the increase relative to baseline. Abbreviations: VE, vaccine effectiveness.

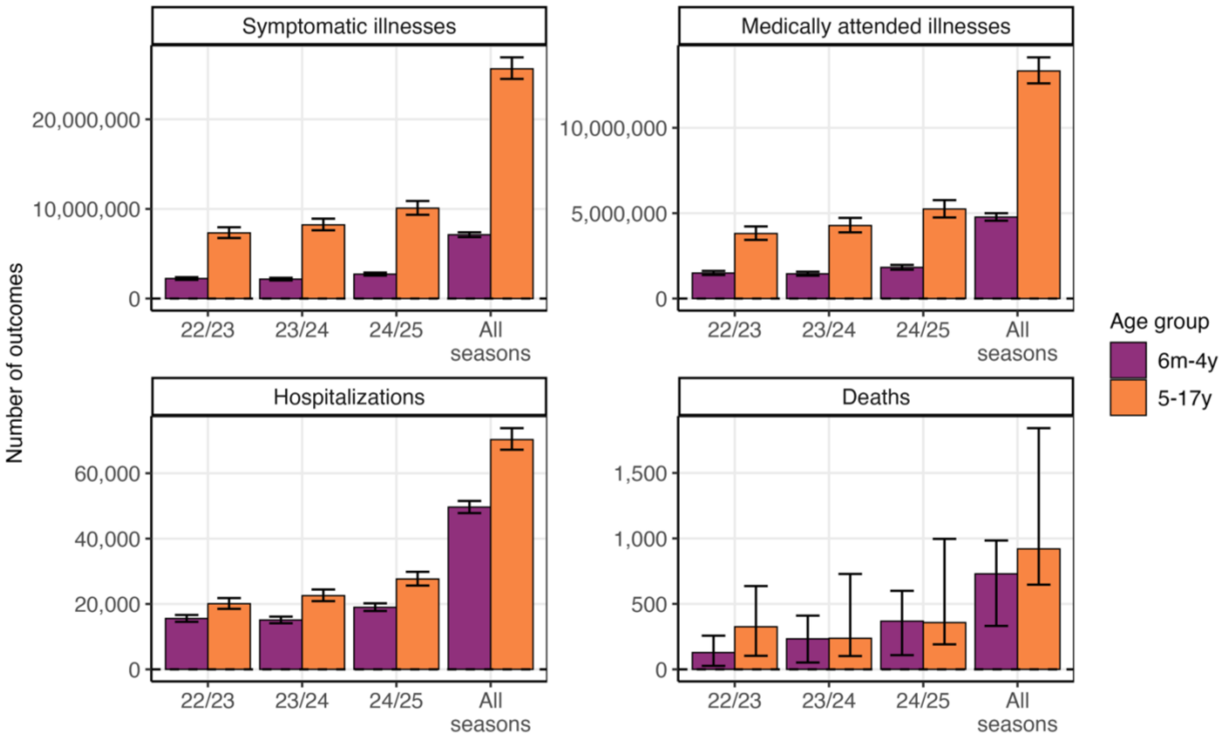


**Figure S5. Estimated number of influenza-associated outcomes that occurred in the baseline scenario.** Bars represent point estimates and error bars are the 95th uncertainty intervals.

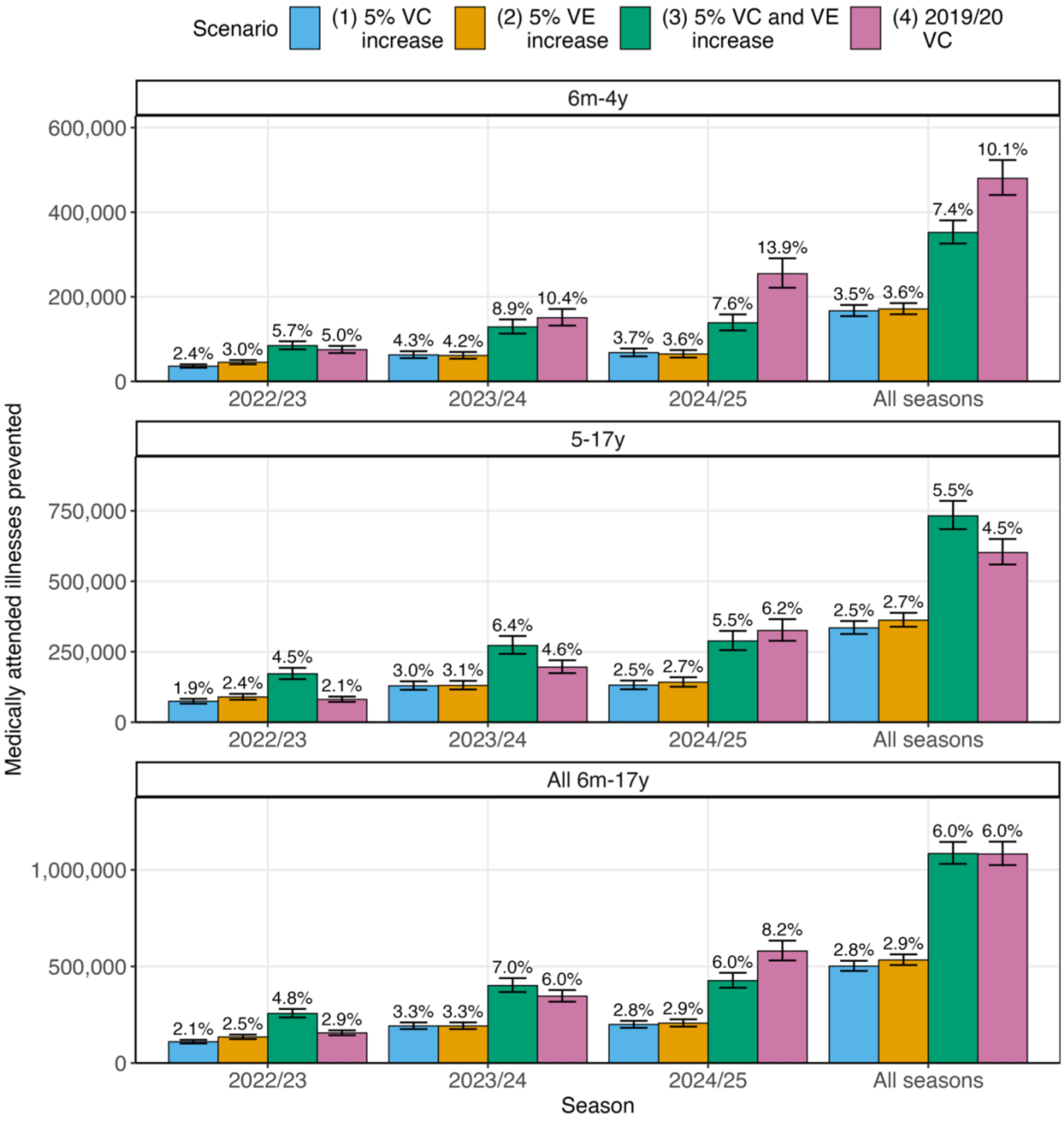


**Figure S6. Estimated influenza-associated medically attended illnesses prevented by vaccination in different vaccination scenarios.** Bars represent point estimates and error bars reflect 95th uncertainty intervals. Numbers above bars are point estimates of the percentage reduction in medically attended illnesses relative to baseline. Abbreviations: VC, vaccination coverage; VE, vaccine effectiveness.

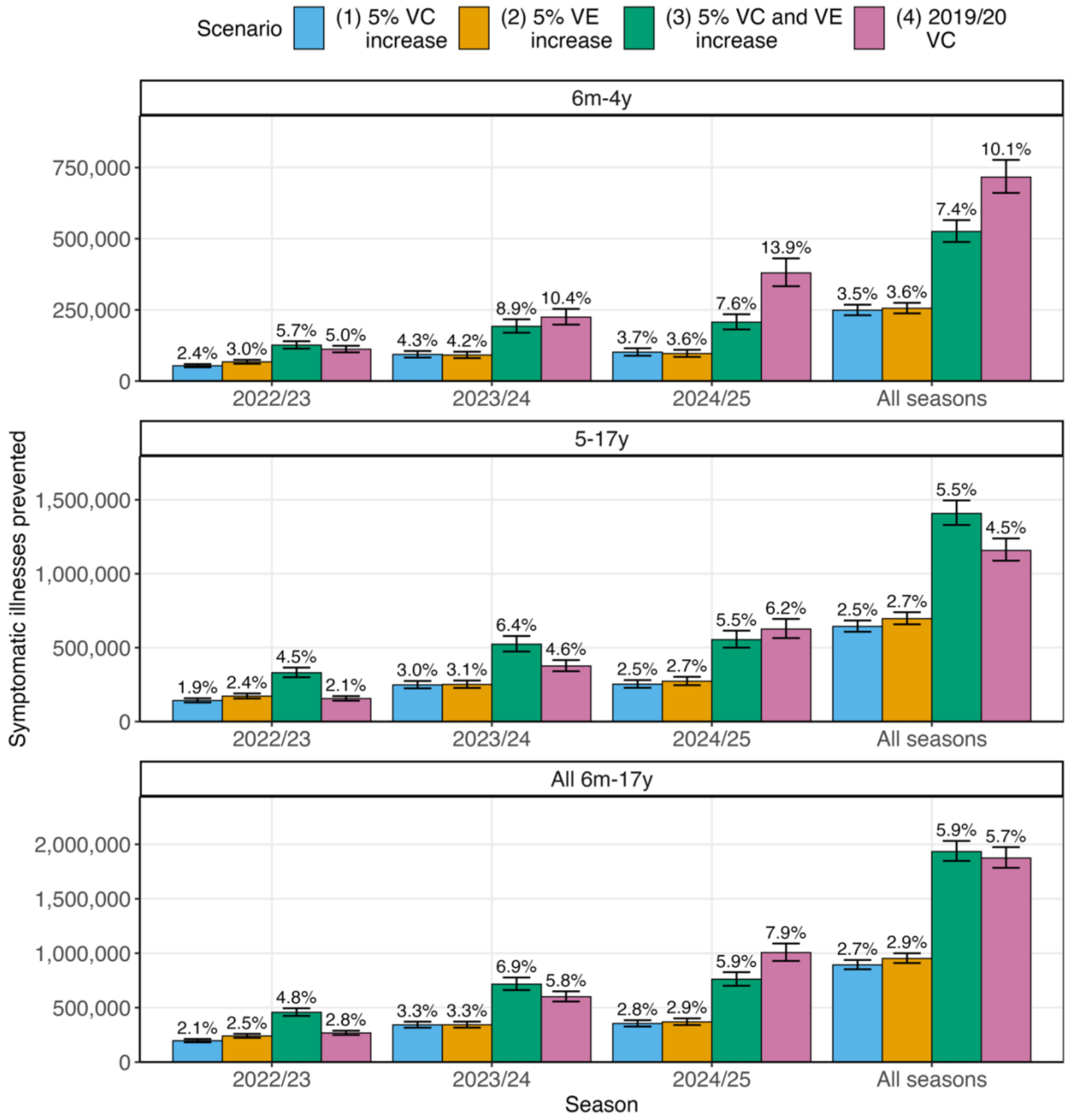


**Figure S7. Estimated influenza-associated symptomatic illnesses prevented by different vaccination scenarios.** Bars represent point estimates and error bars reflect 95th uncertainty intervals. Numbers above bars are point estimates of the percentage reduction in symptomatic illnesses relative to baseline. Abbreviations: VC, vaccination coverage; VE, vaccine effectiveness.

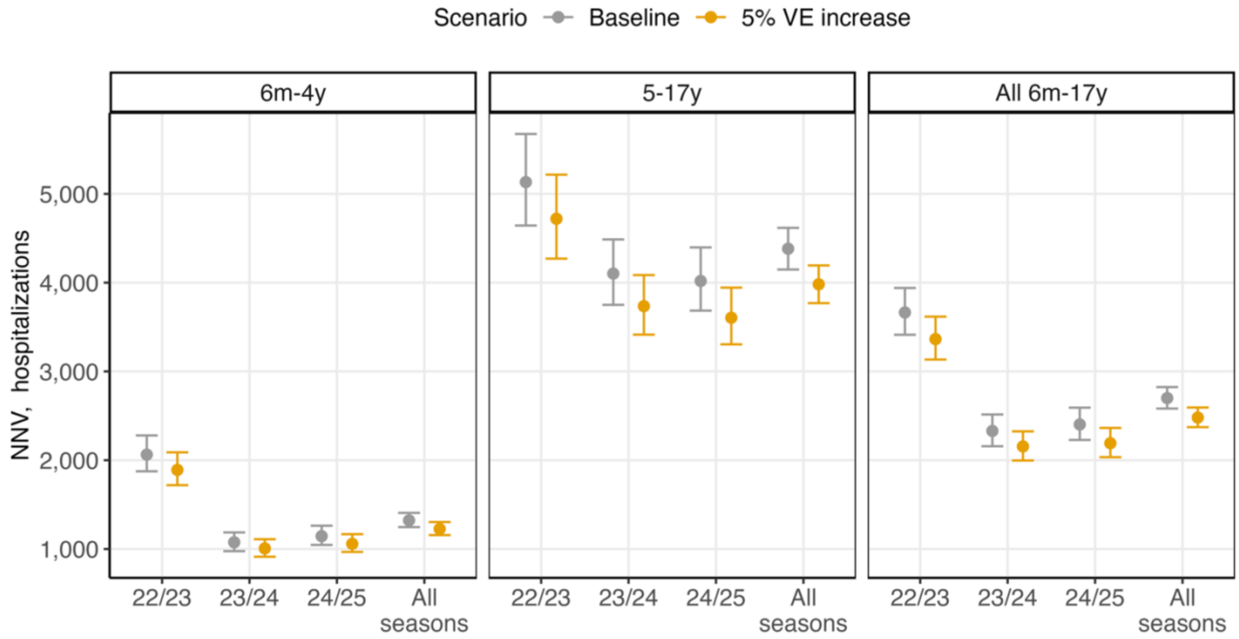


**Figure S8. Number needed to vaccinate (NNV) to prevent one influenza-associated hospitalization.** Circles represent point estimates and error bars reflect 95th uncertainty intervals. Abbreviations: VE, vaccine effectiveness.

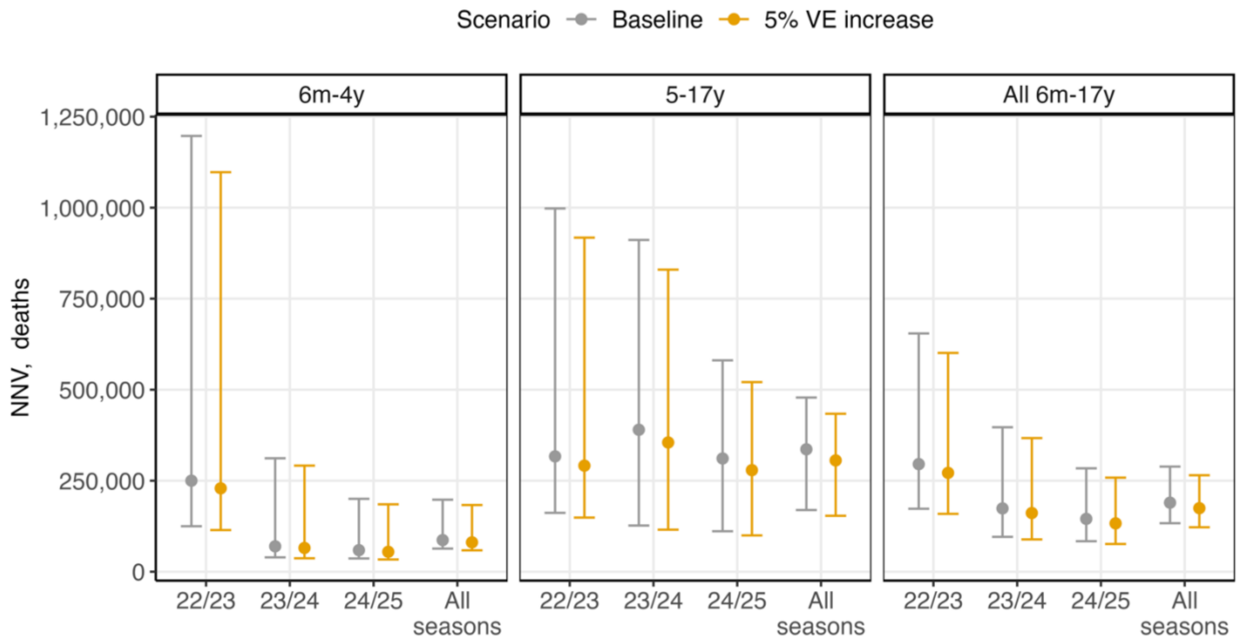


**Figure S9. Number needed to vaccinate (NNV) to prevent one influenza-associated death.** Circles represent point estimates and error bars reflect 95th uncertainty intervals. Abbreviations: VE, vaccine effectiveness.

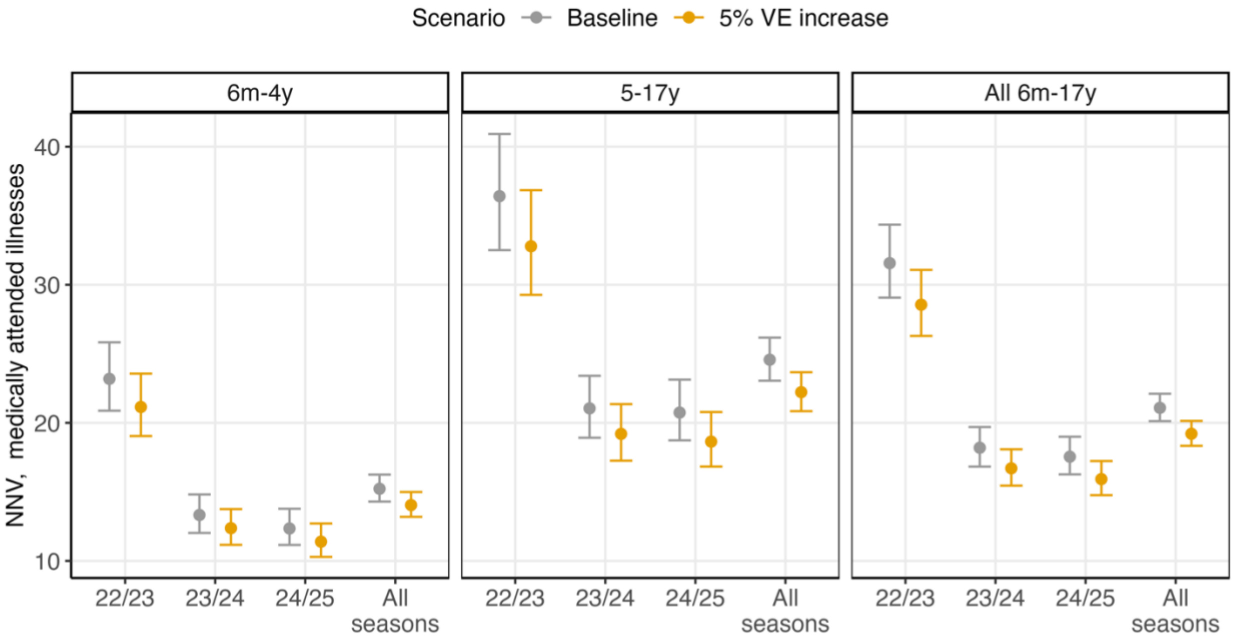


**Figure S10. Number needed to vaccinate (NNV) to prevent one case of influenza-associated medically attended illness.** Circles represent point estimates and error bars reflect 95th uncertainty intervals. Abbreviations: VE, vaccine effectiveness.

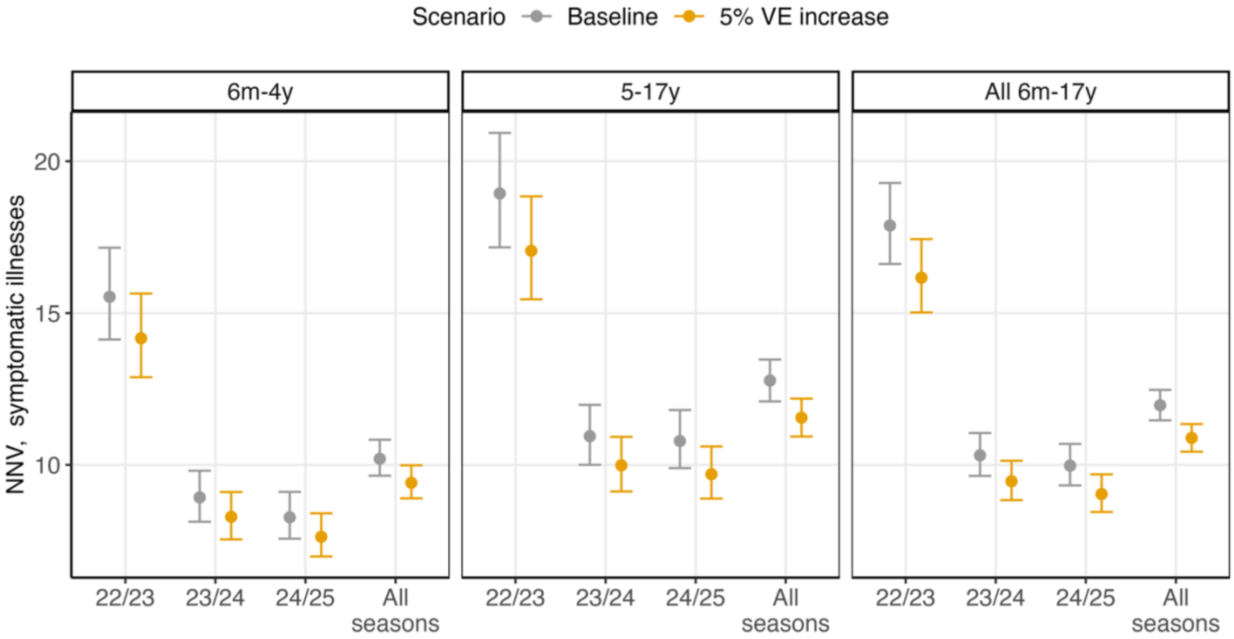


**Figure S11. Number needed to vaccinate (NNV) to prevent one case of influenza-associated symptomatic illness.** Circles represent point estimates and error bars reflect 95th uncertainty intervals. Abbreviations: VE, vaccine effectiveness.
